# Intrapartum Oxytocin and Maternal Outcomes Following Vaginal and Unscheduled Cesarean Delivery

**DOI:** 10.64898/2026.06.20.26356155

**Authors:** Hadas Allouche-Kam, Isha H. Arora, Mary C. Lee, Jiajia Zhang, Francine Hughes, Sharon Dekel

## Abstract

**Objective:** To examine whether intrapartum synthetic oxytocin exposure for labor induction or augmentation is associated with breastfeeding and postpartum depressive and traumatic stress symptoms.

**Methods:** We studied 1,296 postpartum women who delivered at a single tertiary care center, with assessments from the third trimester through approximately two months postpartum. Intrapartum oxytocin exposure was obtained from electronic medical records. Outcomes included exclusive breastfeeding, postpartum depression, and childbirth-related traumatic stress. Analyses were stratified by delivery mode and adjusted for key maternal and obstetric covariates.

**Results:** Overall, 63.3% of participants received intrapartum oxytocin. Among participants with vaginal delivery, oxytocin exposure was associated with lower exclusive breastfeeding at two months after adjustment (58.2% vs 70.3%; adjusted RR 0.86, 95% CI 0.76–0.97; *p* = 0.02), but not with postpartum mental health outcomes. Among participants with unscheduled cesarean delivery, oxytocin exposure was independently associated with higher immediate postpartum depressive symptoms (*F* = 4.97, *p* = 0.03), acute childbirth-related stress (*F* = 4.56, *p* = 0.03), and two-month childbirth-related posttraumatic stress symptoms (*F* = 4.30, *p* = 0.04), but not two-month depressive symptoms.

**Conclusion:** Intrapartum oxytocin exposure was associated with lower exclusive breastfeeding after vaginal delivery and modestly higher childbirth-related distress after unscheduled cesarean delivery. These findings suggest that oxytocin exposure may mark or contribute to postpartum vulnerability in specific delivery contexts.

**Précis:** Intrapartum synthetic oxytocin exposure was associated with lower exclusive breastfeeding after vaginal delivery and modestly higher childbirth-related distress after unscheduled cesarean delivery.

## Introduction

Intrapartum exposure to synthetic oxytocin (OXT) is among the most common pharmacologic interventions in obstetric practice and is widely used for labor induction and augmentation. Nearly half of all births in the United States involve labor induction or augmentation.^1^ Clinical studies show that intrapartum OXT is associated with shorter labor duration and reduced maternal infectious morbidity without increased maternal or neonatal adverse outcomes.^2,3^

An important question concerns the implications of intrapartum OXT exposure for postpartum maternal adjustment beyond physical health. Endogenous OXT regulates milk letdown, stress responses and maternal behavior,^4–7^ raising the possibility that exogenous OXT may influence postpartum recovery trajectories^8^. Existing studies examining OXT exposure and breastfeeding outcomes have produced mixed findings, with some reporting lower exclusive breastfeeding rates among exposed patients and others finding no meaningful associations.^4,5,9–13^

The relationship between intrapartum OXT exposure and postpartum mental health remains less understood. Maternal mental health conditions are leading contributors to pregnancy-associated morbidity and mortality and adverse child outcomes.^14,15^ Although reviews report insufficient evidence linking OXT exposure with postpartum depression,^16,17^ little is known about associations with childbirth-related traumatic stress. Because childbirth can be experienced as traumatic^18–21^ and OXT signaling has been implicated in stress-related processes,^7,22^ this association warrants investigation.

We therefore examined intrapartum OXT exposure as a potential correlate of postpartum maternal adjustment, focusing on breastfeeding, depression, and childbirth-related traumatic stress. Because the clinical implications of OXT exposure may differ by birth context, we evaluated associations separately for vaginal delivery and unscheduled cesarean delivery.

## Materials and Methods

### Study Sample

The study sample included 1,296 participants who delivered at a large academic tertiary care center and enrolled from the third trimester. They were recruited during routine outpatient perinatal visits and through the hospital’s Patient Gateway portal. Among them, 820 participants (63.3%) received intrapartum synthetic OXT for labor induction and/or augmentation, and 476 (36.7%) did not receive intrapartum OXT based on electronic medical records (EMRs).

This study is part of a larger research project concerning childbirth experience and maternal mental health.^18^ Exclusion criteria were maternal age < 18, severe mental illness or substance abuse, and stillbirth per EMR. Additional exclusions for this study included scheduled cesarean delivery and multifetal gestation.

Participants were studied during pregnancy, on average, gestational age was 35.16 weeks (SD = 3.37), in the first days after childbirth, primarily during maternity hospitalization stay, on average 41.40 hours postpartum (SD = 52.32), and again at ∼2 months postpartum (M = 1.95 months, SD = 0.36).

Participants were enrolled between October 2016 and October 2022 (Cohort 1; excluding patients who delivered during the COVID-19 pandemic) and from March 2023 with data available for patients delivering through mid-December 2025 (Cohort 2). Participants provided implied consent by responding to study questionnaires after receiving information about the study. The research project was approved by the hospital’s Human Research Committee.

### Study Measures

Breastfeeding status was assessed at around two months postpartum using the Index of Breastfeeding Status (IBS), which classifies infant feeding as exclusive breastfeeding, mixed breastfeeding (with supplement), stopped breastfeeding, or never breastfed.

Mental health symptoms were assessed repeatedly for maternal depression and traumatic stress. Depression was assessed during pregnancy and in the postpartum assessments using the Edinburgh Postnatal Depression Scale (EPDS).^23^ This is the widely implemented screening 10-item screener for perinatal depression in clinical practice.^24^ It measures symptom endorsement over the last 7 days, with higher scores indicating greater severity. EPDS scores for pregnancy and the second postpartum time point were primarily derived from EMR data. Internal consistency was good (Cronbach’s α = 0.83).

Acute distress during childbirth or immediately after was assessed with the Peritraumatic Distress Inventory (PDI)^25^. It entails 13-item targeting emotional and physiological distress in regard to a specified event (here, childbirth), with higher scores indicating greater distress. The PDI has been used in perinatal samples and shows good validity in identifying cases of childbirth-related posttraumatic stress disorder^18,19^ (α = 0.84).

Posttraumatic stress disorder symptoms regarding recent childbirth were measured at two months postpartum using the PTSD Checklist for DSM-5 (PCL-5),^26^ the recommended patient report tool by the Veterans Affairs. It has 20-items corresponding to the DSM-5 PTSD symptoms, with higher scores reflecting greater symptom severity in the past month. The PCL-5 demonstrates strong psychometric properties and has been validated for the postpartum population against diagnostic assessment^27^ (α = 0.91).

Obstetrical and childbirth-related data were extracted from EMR. This included intrapartum OXT exposure, parity, gestational week, mode of delivery, labor and delivery complications (e.g., hypertensive disorders of pregnancy, hemorrhage, shoulder dystocia, manual removal of the placenta, fetal intolerance of labor, etc.), neuraxial anesthesia (i.e., epidural and spinal/combined spinal-epidural anesthesia), and neonatal intensive care unit (NICU) admission. Demographic (i.e., maternal age, marital status, education, and income), medical (mental health history), and antepartum breastfeeding intention information were obtained using single-item questions or EMR.

## Data Analysis

All analyses were conducted in R (version 4.5.2)^28^ using packages for latent variable modeling, robust standard errors, and model diagnostics. Intrapartum OXT exposure was examined as a binary variable (exposure versus non-exposure).

Missing data for outcome variables and variables treated as covariates accounted for 11.34% of the dataset and were assumed to be missing at random (MAR). Analyses were conducted using available data relevant to each model and stratified by delivery mode.

To examine the association between intrapartum OXT (exposure versus no exposure) and breastfeeding (exclusive versus all other categories), we performed Chi-squared analysis followed by relative risk (RR). Next, adjusted associations were estimated using generalized linear models with a Poisson distribution and log link to estimate risk ratios (RRs), including relevant obstetric and clinical covariates, i.e., parity, neuraxial anesthesia, labor and delivery complications, all which were associated with OXT exposure (defined a-priori, p < 0.10) and antepartum breastfeed intention, which was associated with postpartum breastfeeding status (p < 0.001). Additionally, assisted vaginal (yes versus no) was treated as a covariate in the vaginal delivery analyses.

To examine the association between intrapartum OXT and mental health outcomes (depression and traumatic stress), we performed independent t-tests followed by adjusted analyses using analysis of covariance (ANCOVA) models implemented through linear regression and evaluated using the F statistic. Covariates included the variables noted above (excluding breastfeeding intention) and antepartum depression when examining postpartum depression outcomes.

Sensitivity analyses examined differences between OXT exposure groups (OXT use from the time of labor induction versus augmentation only) across study outcomes.

## Results

The mean maternal age of the sample was 33.54 (SD = 4.01) years, and 56.17% (n = 728) of participants were primiparous. Most participants (95.76%, n = 1,241) delivered at term. In total, 81.48% (n = 1,056) delivered vaginally. 18.52% (n = 240) had unscheduled cesarean delivery. Table 1 presents demographic, obstetric, and clinical characteristics for OXT exposure by mode of delivery.

**Table 1.**
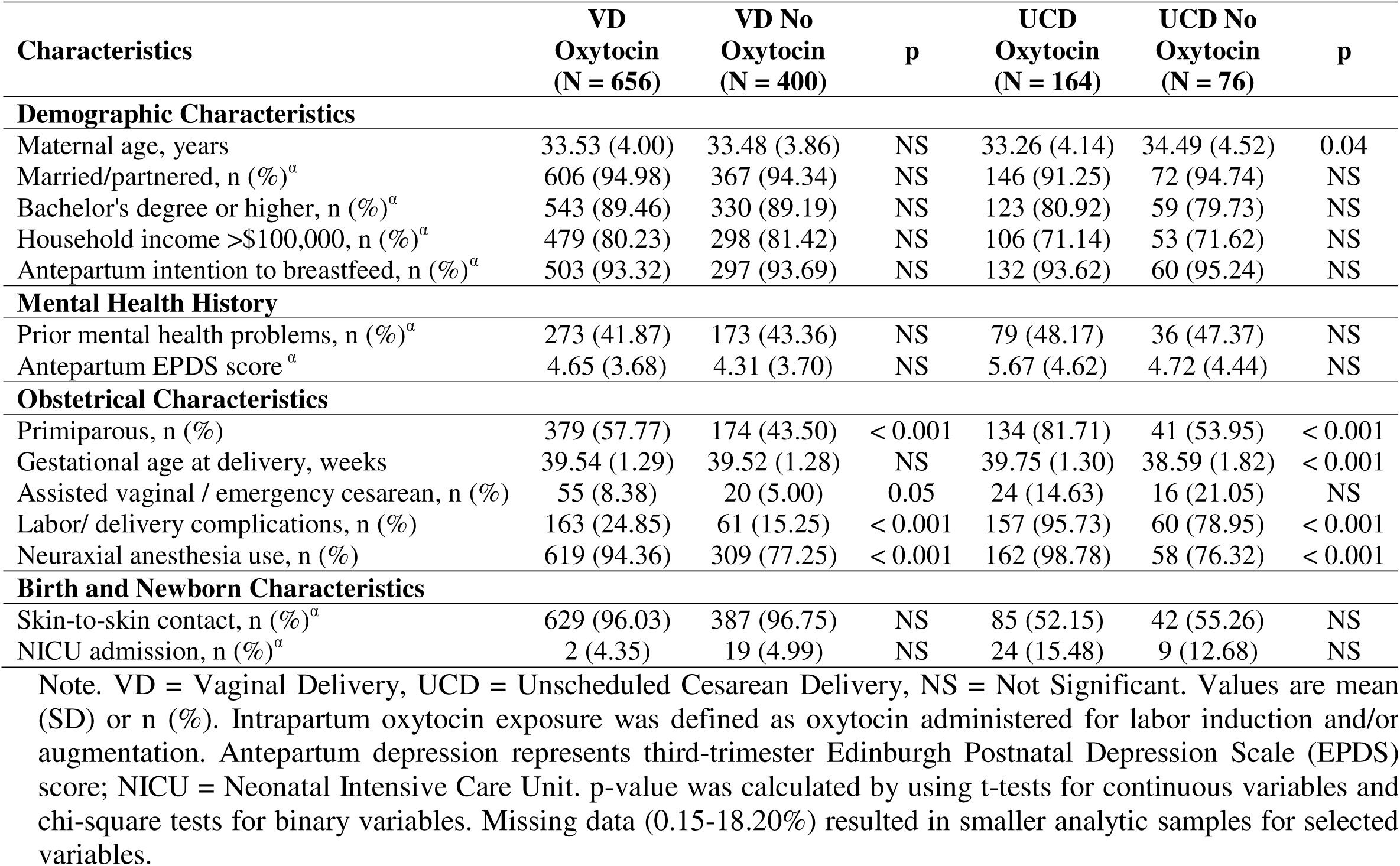
Baseline characteristics by intrapartum oxytocin exposure and delivery mode.

### Intrapartum OXT Exposure and Vaginal Delivery

Among participants who had vaginal delivery, 58.25% (n = 233) of those exposed to OXT reported exclusive breastfeeding at two months postpartum, compared with 70.28% (n = 175) of those not exposed, yielding a *RR* of 0.83 (95% *CI* 0.74 - 0.93, *p* = 0.001). The increased risk of not exclusively breastfeeding associated with intrapartum OXT remained significant after adjustment for study covariates, with an adjusted *RR* of 0.86 (95% *CI* 0.76 - 0.97, *p* = 0.02) (Figure 1). Exclusive breastfeeding rates did not differ between participants exposed to continuous OXT from the time of induction and those exposed for augmentation alone (Supplementary Table 1).

**Figure 1.**
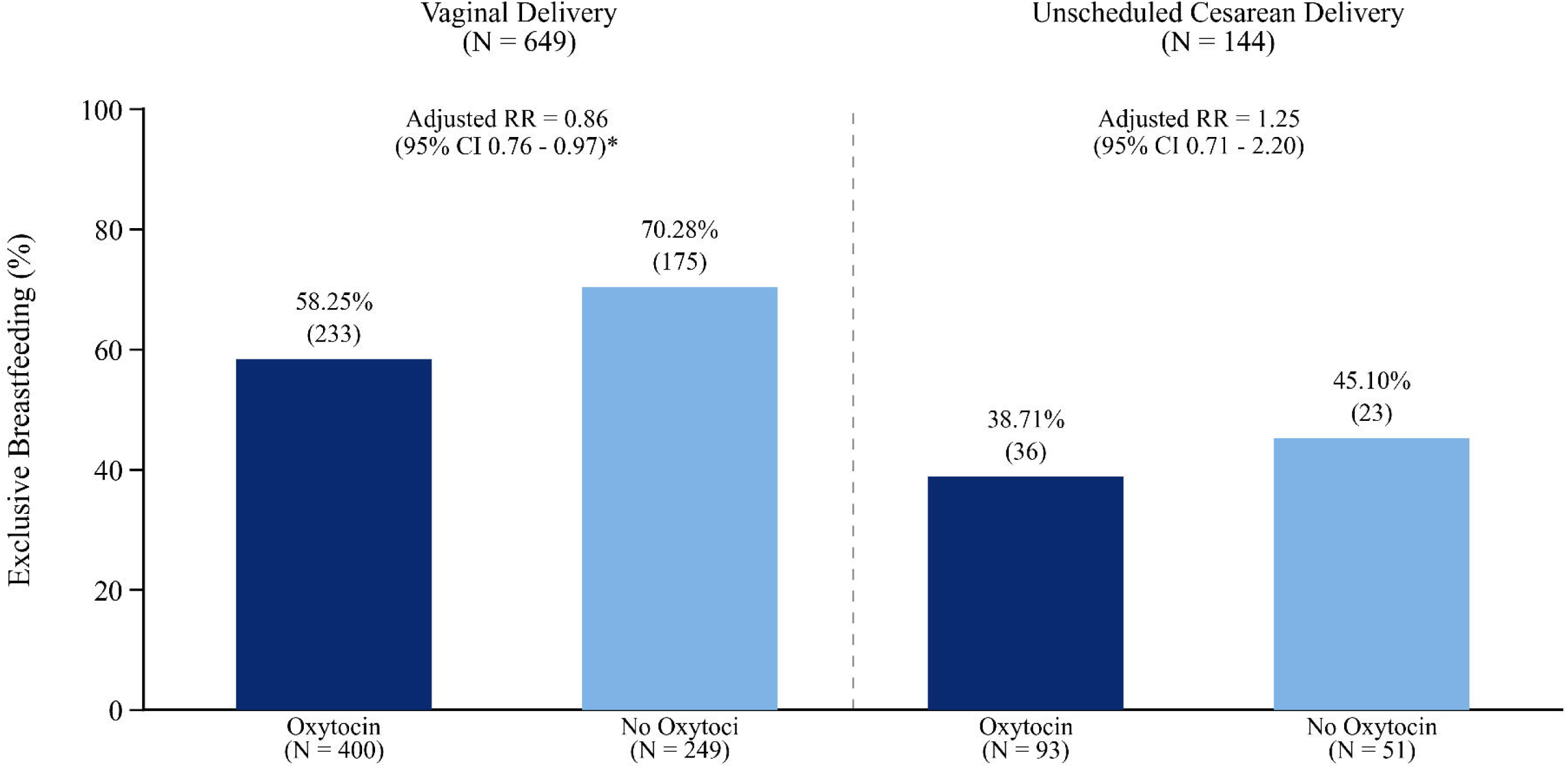
Breastfeeding by intrapartum oxytocin exposure and delivery mode. The figure depicts rates of exclusive breastfeeding at the two months postpartum assessment for vaginal and unscheduled cesarean deliveries separately, for intrapartum oxytocin exposure (induction and/or augmentation) in comparison to no exposure. RR = Relative Risk. Adjusted model controls for study covariates. * p<0.05.

Among the vaginal delivery cohort, acute stress levels during childbirth and posttraumatic stress disorder symptom severity at two months postpartum were low and were not related to OXT exposure, adjusted model, *F* = 0.07, *p* = 0.80 and *F* = 1.21, *p* = 0.27, respectively (Table 2). Similarly, depressive symptom levels in the immediate postpartum and at two months postpartum did not differ by intrapartum OXT exposure, adjusted, *F* = 1.16, *p* = 0.28 and *F* = 2.97, *p* = 0.09, respectively (Table 2). There were also no significant differences in stress or depression levels among OXT exposure groups (Supplementary Table 1).

**Table 2.** Maternal mental health outcomes by oxytocin exposure and delivery mode.

| **Outcome** | **VD**<br>**Oxytocin** | **VD No**<br>**Oxytocin** | **Adjusted F** | **UCD**<br>**Oxytocin** | **UCD No**<br>**Oxytocin** | **Adjusted F** |
| --- | --- | --- | --- | --- | --- | --- |
| **Depression, Immediate PP** | 4.15 (4.11) | 3.59 (3.32) | 1.16 (2.13*) | 5.75 (4.78) | 4.03 (4.25) | 4.97* (2.57*) |
| **Depression, 2-months PP** | 3.85 (3.74) | 3.79 (3.72) | 2.97 (0.19) | 4.51 (4.30) | 4.78 (4.74) | 0.64 (-0.41) |
| **Acute Stress, Immediate PP** | 5.78 (6.43) | 5.78 (6.00) | 0.07 (-0.01) | 12.31 (9.51) | 8.21 (8.59) | 4.56* (3.16**) |
| **PTSD symptoms, 2-months PP** | 4.68 (7.07) | 4.14 (6.41) | 1.21 (1.08) | 8.54 (10.73) | 5.98 (7.79) | 4.30* (1.56) |

Note. VD = Vaginal Delivery, UCD = Unscheduled Cesarean Delivery. PP = Postpartum. Values are mean (SD). Adjusted F values, followed by unadjusted t-test values in parentheses. Adjusted statistics represent ANCOVA models controlling for relevant maternal and obstetrical covariates. Depression was assessed using the Edinburgh Postnatal Depression Scale (EPDS), acute stress using the Peritraumatic Distress Inventory (PDI), and childbirth-related posttraumatic stress disorder symptoms using the PTSD Checklist for DSM-5 (PCL-5). Missing outcome data resulted in smaller analytic samples. \*\* $p < 0.01$ , \* $p < 0.05$ .

### Intrapartum OXT Exposure and Unscheduled Cesarean Delivery

Among participants who underwent unscheduled cesarean delivery, exclusive breastfeeding at around 2-month postpartum was reported by 38.71% (n = 36) of the OXT-exposed, compared with 45.10% (n = 23) of those not exposed, but rates were not significantly different, adjusted *RR* 1.25, 95% *CI* 0.71 - 2.20, *p* = 0.43 (Figure 1). Exclusive breastfeeding rates did not differ within OXT-exposed groups (Supplementary Table 1).

Among the unscheduled cesarean delivery cohort, those exposed to intrapartum OXT reported higher depression and acute stress response to childbirth at the immediate postpartum assessment compared with those not exposed to OXT. For depression, adjusted model, *F* = 4.97, *p* = 0.03 (partial η^2^ = 0.02), and for acute stress, *F* = 4.56, *p* = 0.03 (partial η^2^ = 0.02) (Table 2). The intrapartum OXT exposure group also had higher posttraumatic stress symptoms at the 2 months postpartum assessment, adjusted model, *F* = 4.30, *p* = 0.04 (partial η^2^ = 0.03), although no differences in depression levels (Table 2). Moreover, PTSD symptoms varied by the timing of OXT exposure. Participants exposed to OXT for augmentation alone reported higher symptom levels than those exposed from the time of induction (adjusted, *F* = 4.12, *p* = 0.05) (Supplementary Table 1).

## Discussion

The primary aim of this study was to examine whether intrapartum OXT exposure is associated with postpartum breastfeeding and maternal psychological outcomes across different delivery contexts. Distinguishing between vaginal and unscheduled cesarean deliveries is important because these represent clinically distinct birth experiences, with unscheduled cesarean delivery often reflecting greater obstetric complexity and a higher-risk context for adverse postpartum mental health outcomes.^18,29^

Our findings suggest that the implications of intrapartum OXT exposure may depend on delivery context. Among participants with vaginal delivery, OXT exposure was associated with a lower likelihood of exclusive breastfeeding in the early postpartum period but was not associated with maternal mental health outcomes. In contrast, among participants with unscheduled cesarean delivery, OXT exposure was associated with higher levels of depressive symptoms, acute childbirth-related distress, and subsequent childbirth-related traumatic stress, after accounting for parity, neuraxial anesthesia use, labor and delivery complications. Although effect sizes were small, these findings suggest that OXT exposure may contribute additional risk in a delivery context already characterized by elevated vulnerability to postpartum psychological distress. However, the observational nature of the study precludes causal inference, and underlying obstetric acuity, prolonged labor, or other clinical factors associated with OXT use may explain the observed associations.

Understanding whether obstetrical interventions influence breastfeeding has important clinical implications. Exclusive breastfeeding remains a public health priority, with recommendations for exclusive breastfeeding through 6 months and continued breastfeeding through 1 year.^30^ Among participants with vaginal delivery, exclusive breastfeeding was relatively high in the absence of intrapartum OXT exposure, approaching 70%, but was lower among OXT-exposed participants, with 58% exclusively breastfeeding at 2 months postpartum. This finding is notable because most participants in the vaginal delivery subgroup did not experience major birth complications. In contrast, exclusive breastfeeding rates were low among participants with unscheduled cesarean delivery regardless of OXT exposure, suggesting that broader obstetric and clinical factors may be more influential in this medically complex subgroup.

Research on the implications of intrapartum OXT exposure for maternal mental health remains limited. To our knowledge, this study is the first to examine acute childbirth-related stress in relation to OXT exposure, particularly in the context of unscheduled cesarean delivery. In this subgroup, OXT exposure was associated with persistent traumatic stress in response to childbirth but not depressive symptoms. This pattern suggests that OXT exposure may be more closely related to the traumatic appraisal of childbirth than to postpartum mood symptoms.

Prior work has shown that traumatic stress symptoms are more common after unscheduled cesarean delivery than after vaginal delivery.^18,29^ Preclinical studies suggest that, in stressful contexts, OXT may heighten threat processing and strengthen traumatic memory consolidation by increasing the salience of emotional cues^7,8,22^ In the present study, childbirth-related traumatic stress symptoms were also higher when OXT was administered for labor augmentation only than when exposure began with induction. This pattern may reflect the stressful and unexpected escalation from attempted vaginal delivery to unscheduled cesarean delivery rather than prolonged OXT exposure per se, underscoring the importance of interpreting OXT exposure within its broader obstetric context.

This study has several strengths. It included a large cohort of participants delivering at a major academic medical center, with data collected from pregnancy through the postpartum period. The study assessed both breastfeeding and maternal mental health outcomes, including childbirth-related traumatic stress, and used electronic medical record data to account for key obstetric and clinical factors.

Several limitations should be noted. The observational design precludes causal inference, and unmeasured factors may have influenced both intrapartum OXT use and postpartum outcomes. Because postpartum OXT for hemorrhage prophylaxis is part of standard obstetric care, differences attributable to intrapartum OXT exposure may have been attenuated. Endogenous OXT levels were not measured, and maternal outcomes were assessed using validated self-report instruments rather than clinical interviews. Participants who completed both postpartum assessments had similar obstetric characteristics to non-completers but differed in demographic background, which may limit generalizability.

In conclusion, intrapartum OXT exposure was associated with lower exclusive breastfeeding after vaginal delivery and modestly higher childbirth-related distress after unscheduled cesarean delivery. These findings suggest that OXT exposure may mark or contribute to postpartum vulnerability in specific delivery contexts. Further research is needed to clarify the mechanisms underlying these associations and their implications for postpartum health.

## Supporting information

Supplementary Table S1. Breastfeeding and maternal mental health outcomes by oxytocin indication and delivery mode.

## Data Availability

All data produced in the present study are available upon reasonable request to the authors

## Trial registration number

Not applicable.

## Prior presentation at a scientific meeting

None.

## Disclaimer

Not applicable.

## Acknowledgments

We thank Christina Pham, Eunice Chon, Onyeka Agwu, and Sabrina J. Chan for their assistance with electronic medical records data extraction.

## Declaration of generative-AI use

During the preparation of this work, the authors used ChatGPT-5.5 to improve language and readability. The authors reviewed and edited the content as needed and take full responsibility for the final version.

