## Supplementary Table S1. Breastfeeding and maternal mental health outcomes by oxytocin indication and delivery mode. for "Intrapartum Oxytocin and Maternal Outcomes Following Vaginal and Unscheduled Cesarean Delivery"

| Outcome | **VD Induction** | **VD Augmentation** | **Adjusted F/RR** | **UCD Induction** | **UCD Augmentation** | **Adjusted F/RR** |
| --- | --- | --- | --- | --- | --- | --- |
| **Exclusive Breastfeeding, 2-months PP** | 122 (57.01) | 111 (59.68) | 1.05  (0.89-1.24) | 18 (40.00) | 18 (37.50) | 0.84  (0.49-1.44) |
| **Depression, Immediate PP** | 3.90 (4.01) | 4.41 (4.20) | 1.49 | 5.80 (5.08) | 5.71 (4.47) | 1.74 |
| **Depression, 2-months PP** | 3.80 (3.75) | 3.90 (3.74) | 0.01 | 4.39 (4.29) | 4.64 (4.35) | 1.64 |
| **Acute Stress, Immediate PP** | 5.41 (5.84) | 6.15 (6.98) | 1.17 | 12.21 (9.09) | 12.42 (9.97) | 0.10 |
| **PTSD symptoms, 2-months PP** | 4.85 (6.94) | 4.50 (7.20) | 0.24 | 6.45 (7.14) | 10.45 (12.96) | 4.12* |

Note. VD = Vaginal Delivery, UCD = Unscheduled Cesarean Delivery, PP = Postpartum, RR = Risk Ratio. Values are mean (SD). Adjusted statistics represent ANCOVA models controlling for relevant maternal and obstetrical covariates. Exclusive breastfeeding measured in comparison to no exclusive breastfeeding (mix feeding, stopped or never breastmilk). Depression was assessed using the Edinburgh Postnatal Depression Scale (EPDS), acute stress using the Peritraumatic Distress Inventory (PDI), and childbirth-related posttraumatic stress symptoms using the PTSD Checklist for DSM-5 (PCL-5). Missing outcome data resulted in smaller analytic samples. *p < 0.05.
